# The Impact of Insulin Resistance on Grey Matter Changes Along the Alzheimer’s Disease Continuum Insulin Resistance and Grey Matter in AD

**DOI:** 10.1101/2025.11.17.25339566

**Authors:** Valentina Bettonagli, Alice Galli, Elena Bazzoli, Chiara Tolassi, Salvatore Caratozzolo, Bianca Gumina, Ilenia Libri, Daniel Ferreira, Tiago Fleming Outeiro, Andrea Pilotto, Alessandro Padovani, the Alzheimer’s Disease Neuroimaging Initiative

**Affiliations:** Neurology Unit, Department of Clinical and Experimental Sciences, University of Brescia, Italy; Department of continuity of care and frailty, Neurology Unit, ASST Spedali Civili Hospital, Brescia, Italy; Neurobiorepository and Laboratory of Advanced Biological Markers, University of Brescia and ASST Spedali Civili Hospital, Brescia, Italy; Laboratory of Digital Neurology and Biosensors, University of Brescia, Italy; Nutri Neuro Med, Desenzano del Garda, Italy; Department of Neurobiology, Care Sciences and Society (NVS) Center for Alzheimer Research, Division of Clinical Geriatrics, Karolinska Institutet, Stockholm, Sweden; Facultad de Ciencias de la Salud, Universidad Fernando Pessoa Canarias, Las Palmas, España; University Medical Center Goettingen, Department of Experimental Neurodegeneration, Center for Biostructural Imaging of Neurodegeneration, Goettingen, Germany; Translational and Clinical Research Institute, Faculty of Medical Sciences, Newcastle University, Framlington Place, Newcastle Upon Tyne, NE2 4HH, UK; Brain Health Center, University of Brescia, Italy

## Abstract

**Background and Objectives:** Insulin resistance is emerging as a modifiable risk factor for Alzheimer’s, though its impact on grey matter volume across clinical stages remains poorly understood. The objective of the research is to investigate how insulin resistance affects grey matter integrity across the Alzheimer’s disease continuum using structural MRI.

**Methods:** Imaging, clinical, and metabolic data were extracted from 374 non-diabetic participants within the Alzheimer’s Disease Neuroimaging Initiative (ADNI) dataset. Participants were classified as cognitively impaired (CI: n=186; 137 mild cognitive impairment, 49 early-to-moderate dementia; all AD biomarker positive) or cognitively unimpaired (CU: n=188; 122 amyloid-negative, 66 amyloid-positive). Insulin resistance was assessed at the time of MRI and clinical evaluation using the dichotomized triglyceride-glucose index (TyG). The Interactions between TyG and diagnostic group on grey matter volume were investigated using both voxel-wise and region-of-interest (ROI) based analyses, adjusted for age, sex, education, vascular risk factors, and global cognitive performance across the AD continuum.

**Results:** Insulin resistance significantly impacted gray matter volume across the AD continuum, demonstrating stage-dependent effects. In early AD disease stages, insulin resistance was associated with lower grey matter volume in fronto-parietal regions, a finding that extended to several cortical areas in CI individuals. Temporal and fronto-limbic regions were particularly highlighted by the IR-diagnosis interaction. In amyloid-positive CU individuals, IR was linked to bilateral temporal atrophy, in contrast to amyloid-negative CU participants.

**Discussion:** This study underscores the impact of insulin resistance on brain structure across the AD continuum, particularly within key vulnerability areas characteristic of AD pathology. These findings highlight the need for future research into potential therapeutic strategies targeting insulin signaling to mitigate neurodegeneration in AD.

## Introduction

Alzheimer’s disease (AD) is the leading cause of dementia worldwide and is characterized by the accumulation of amyloid-β (Aβ) plaques and neurofibrillary tangles composed of hyperphosphorylated tau protein^1^. While non-modifiable risk factors such as age, sex, and genetic predisposition significantly contribute to AD pathogenesis, increasing attention has been directed toward modifiable factors, including hypercholesterolemia, hypertension, diabetes, and other metabolic changes^2,3^. Among these, insulin resistance (IR) has emerged as a key metabolic factor potentially involved in AD pathophysiology. IR impairs insulin signaling pathways in the brain, which may promote Aβ and tau accumulation, enhance oxidative stress, and increase protein glycation^4–6^. Additionally, IR has been associated with reduced neuronal glucose uptake and utilization, further compromising neuronal function^7^.

Neuroimaging studies in cognitively healthy older adults have demonstrated that IR is associated with structural brain alterations, particularly grey matter atrophy in regions vulnerable to early AD pathology such as the medial temporal lobe, prefrontal cortex, and cingulate cortex^8,9^, along with microstructural abnormalities in white matter tracts^10,11^. Importantly, similar findings have been reported in non-diabetic AD patients, supporting a link between metabolic dysfunction and neurodegeneration independent of overt diabetes^12^.

Recent studies have also highlighted the mechanism linking IR and the AD genetic risk factor *APOE* ε4, potentially disrupting blood-brain barrier integrity and thus potentially facilitating the entry of neurotoxic agents into the brain^13^. These observations emphasize the complex interplay between metabolic dysfunction, individual genetic and somatic susceptibility in driving the pathophysiological cascade of AD.

Despite these advances, the relationship between IR and grey matter volume reduction across the AD continuum remains insufficiently characterized. Specifically, no studies evaluated the relationship between IR and brain structural changes at different clinical stages of AD accounting for genetic risk, clinical variables, metabolic comorbidities, and biomarkers of AD pathology^14–16^. This gap has limited our understanding of how IR may interact with core AD mechanisms to influence brain structure and disease progression.

In this study, we investigate the association between IR and grey matter volume in elderly individuals across the AD continuum using structural MRI data. Leveraging the Alzheimer’s Disease Neuroimaging Initiative (ADNI) cohort, we aim to clarify the contribution of IR to brain atrophy in the context of AD pathology, considering key risk and protective factors and stratifying participants by clinical stage of the disease.

## Methods

### Participants

Participants were retrospectively selected from the Alzheimer’s Disease Neuroimaging Initiative (ADNI) database (adni.loni.usc.edu), including individuals enrolled in the ADNI-1, ADNI-GO, and ADNI-2 phases. The ADNI was launched in 2003 as a public-private partnership, led by Principal Investigator Michael W. Weiner, MD; for up-to-date information, see www.adni-info.org. All participants or their legally authorized representatives provided written informed consent, as documented in ADNI protocols [https://adni.loni.usc.edu/help-faqs/adni-documentation/].

Inclusion criteria were as follows: age between 55 and 90 years; availability of high-resolution 3 Tesla T1-weighted MRI scans; complete apolipoprotein E (*APOE*) genotype data; cerebrospinal fluid (CSF) biomarkers of Alzheimer’s disease (AD) pathology; and comprehensive clinical assessment performed at the time of MRI acquisition, including at least one standardized cognitive measure.

Exclusion criteria included a diagnosis of diabetes mellitus, presence of significant motion artifacts on MRI, extensive white matter hyperintensities, or severe global brain atrophy. All neuroimaging data underwent rigorous quality control to ensure conformity with ADNI imaging standards. The exclusion of individuals with extensive white matter hyperintensities or severe global brain atrophy was based on visual inspection of structural MRI scans by trained raters. This decision was made to reduce the risk of including cases with structural brain alterations that could confound the interpretation of gray matter volume differences related to IR.

Diagnostic classification was based on a combination of cognitive status and CSF amyloid-beta (Aβ) positivity. Aβ positivity (A+) was defined by CSF Aβ42 concentrations below 1030 pg/mL, in accordance with the cut-off value established by Roche for the Elecsys® β-Amyloid (1–42) CSF immunoassay^17,18^. Cognitive status was determined using the Clinical Dementia Rating (CDR) and the Mini-Mental State Examination (MMSE). Participants with CDR = 0 and MMSE > 24 were classified as cognitively unimpaired (CU); those with CDR ≥ 0.5 were considered cognitively impaired (CI), in line with ADNI diagnostic criteria^19^.

The CI group, all of whom were β-amyloid positive, was further divided into individuals with mild cognitive impairment (A+MCI; MMSE > 24) and those with moderate dementia (A+DEM; MMSE ≤ 24)^20^. Cognitively unimpaired participants were also stratified by Aβ status into Aβ-negative A-CU, and Aβ-positive A+CU, in accordance with the cut-off thresholds previously outlined. The A-CU group, which was the only amyloid-negative group, served as the reference population in all comparative analyses, allowing assessment of the specific contributions of amyloid pathology and cognitive impairment to brain gray matter volume.

### Assessment of Insulin Resistance

IR was estimated using the triglyceride-glucose (TyG) index, calculated as: Ln [fasting triglycerides (mg/dL) × fasting glucose (mg/dL) / 2]^21^.

The TyG index has shown good performance in the estimation of IR compared with the HOMA-IR in individuals with and without diabetes while it does not require insulin quantification and it is independent of insulin treatment status^22^.

### MRI Acquisition and Preprocessing

High-resolution 3D T1-weighted MRI scans were acquired using 3 Tesla scanners (Siemens, Philips, GE), utilizing Magnetization-Prepared Rapid Gradient Echo (MPRAGE) and Inversion Recovery Fast Spoiled Gradient Recalled (IR-FSPGR) sequences. Detailed acquisition protocols are publicly available through the ADNI website [https://adni.loni.usc.edu/methods/mri-tool/mri-acquisition/].

Preprocessing was performed using the Computational Anatomy Toolbox (CAT12; http://dbm.neuro.uni-jena.de/cat) within the Statistical Parametric Mapping software (SPM12, Wellcome Centre for Human Neuroimaging, London, UK). Preprocessing steps included spatial normalization to MNI space, segmentation into grey matter, white matter, and CSF, and modulation for nonlinear deformations to preserve local tissue volumes. Images were smoothed using an 8-mm full-width at half maximum Gaussian kernel. Total intracranial volume (TIV) was computed by the CAT12 toolbox as the sum of grey matter, white matter, and cerebrospinal fluid volumes, and included as a covariate in all volumetric analyses. Segmentation accuracy was visually inspected, and data quality was ensured through standardized outlier detection procedures. Identified outliers were excluded from all subsequent statistical analyses^23^.

### Demographics and clinical measures

Baseline demographic data (age, sex, education) and cardio-metabolic variables (namely body mass index [BMI], mean arterial pressure [MAP = DBP + (SBP−DBP)/3]), along with APOE ε4 allele status and MMSE total score (as a measure of global cognitive severity^24^), were included as covariates in the adjusted model due to their known influence on grey matter volume and thickness^25^. CSF biomarker levels (Aβ42, total tau [t-tau], and phosphorylated tau [p-tau]) were collected for descriptive purposes and group characterization.

### Statistical Analysis and MRI

Group differences in demographic, clinical, and CSF biomarker variables were assessed using one-way ANOVA for continuous variables and chi-square tests for categorical variables. Statistical models were designed to evaluate the effects of IR on grey matter volume, accounting for AD pathology, cognitive status, and other relevant factors.

### Voxel-Based Morphometry (VBM)

Whole-brain voxel-based morphometry analyses were performed to investigate the relationship between IR and grey matter volume across diagnostic groups. VBM preprocessing was conducted using grey matter probability maps derived from the tissue segmentation pipeline. All voxel-wise analyses were performed using a threshold of *p* < 0.001 (uncorrected, peak-level), with a minimum cluster extent of 500 contiguous voxels. Significant clusters were anatomically labeled using the AAL3 atlas^26^. All models included covariates for age, sex, education, BMI, MAP, *APOE* ε4 carrier status, TIV, and scan type. Scan type comprised three levels corresponding to the main MRI scanner manufacturers used in ADNI: Philips, Siemens, and GE.

### Diagnosis-Driven Atrophy Mapping

A first multiple regression model was conducted to confirm the effect of diagnostic group (A-CU; A+CU; A+MCI and A+DEM) on grey matter volume across the full sample. This was aimed for the identification of AD-related atrophic regions, to later be used as regions of interest (ROIs) in subsequent analyses.

### IR-by-Diagnosis Interaction

To assess whether the association between IR and grey matter volume differed across clinical groups, a whole-brain VBM model was constructed with an IR × Diagnosis interaction term. This model assessed the differential effect of IR on gray matter volume across clinical stages of disease.

### Subgroup-Specific Analyses adjusted for disease severity

VBM analyses were further conducted within the two major diagnostic classifications, i.e. cognitive impairment group and cognitively unimpaired group, in order to limit additionally adjust for clinical severity and thus limit the impact of disease atrophy on findings. For analyses within the cognitively impaired group, the MMSE score was additionally included as a covariate to account for global cognitive impairment as a proxy for disease severity.

In the CU subgroup, an IR × Amyloid status interaction model was implemented to assess whether the structural effects of IR varied between amyloid-positive and amyloid-negative individuals. This specific analysis focused on cognitively unimpaired subjects was designed in order to evaluate the role of IR in more uniform atrophy subgroup^19^.

### Region-of-Interest (ROI) Analyses

To clarify the directionality and group specificity of VBM findings, mean grey matter volumes were extracted from significant clusters identified in the voxel-wise analyses using CAT12. These were used in post-hoc comparisons to determine whether effects were driven by particular diagnostic groups or consistent across the sample.

Additional hypothesis-driven ROI analyses were conducted on a predefined set of 36 brain regions known to be vulnerable to AD pathology. These ROIs, selected from both the diagnosis-based VBM map and existing literature^27^, were defined using the AAL atlas^26^ and included cortical (e.g., temporal, parietal, insular, and cingulate cortices) and allocortical limbic regions (e.g., hippocampus, amygdala, parahippocampal gyrus, entorhinal cortex). ROI data were analyzed using two-way ANCOVAs with IR and diagnosis as factors. All models included the same covariates as the VBM analyses. To minimize scanner-related variability, ROI values were harmonized across MRI acquisition sites using the ComBat method^28^, a batch-effect correction technique originally developed for genomics and later adapted for neuroimaging.

### Statistical threshold and multiple comparisons correction

Statistical threshold was set at p=0.05, false discovery rate (FDR) correction was applied to control for multiple comparisons across the 36 ROIs and reduce the likelihood of type I errors.

## Results

Out of 1205 participants totally enrolled, 487 elderly individuals met all the selection criteria (Tables 1-3). One-hundred thirteen out of the 487 individuals showed motion artifacts, extensive white matter hyperintensities or poor image quality upon visual and technical inspection, and were subsequently excluded from further analyses. The total sample consisted of 374 participants, including 188 cognitively unimpaired individuals and 186 cognitively impaired individuals. The cognitively unimpaired group was further divided into 122 Aβ-negative A-CU and 66 Aβ-positive A+CU participants, while the cognitively impaired group included 137 CI individuals with mild cognitive impairment A+MCI and 49 individuals with dementia A+DEM^19,20^ (see the flowchart - Supplementary Figure 1).

**Table 1.**
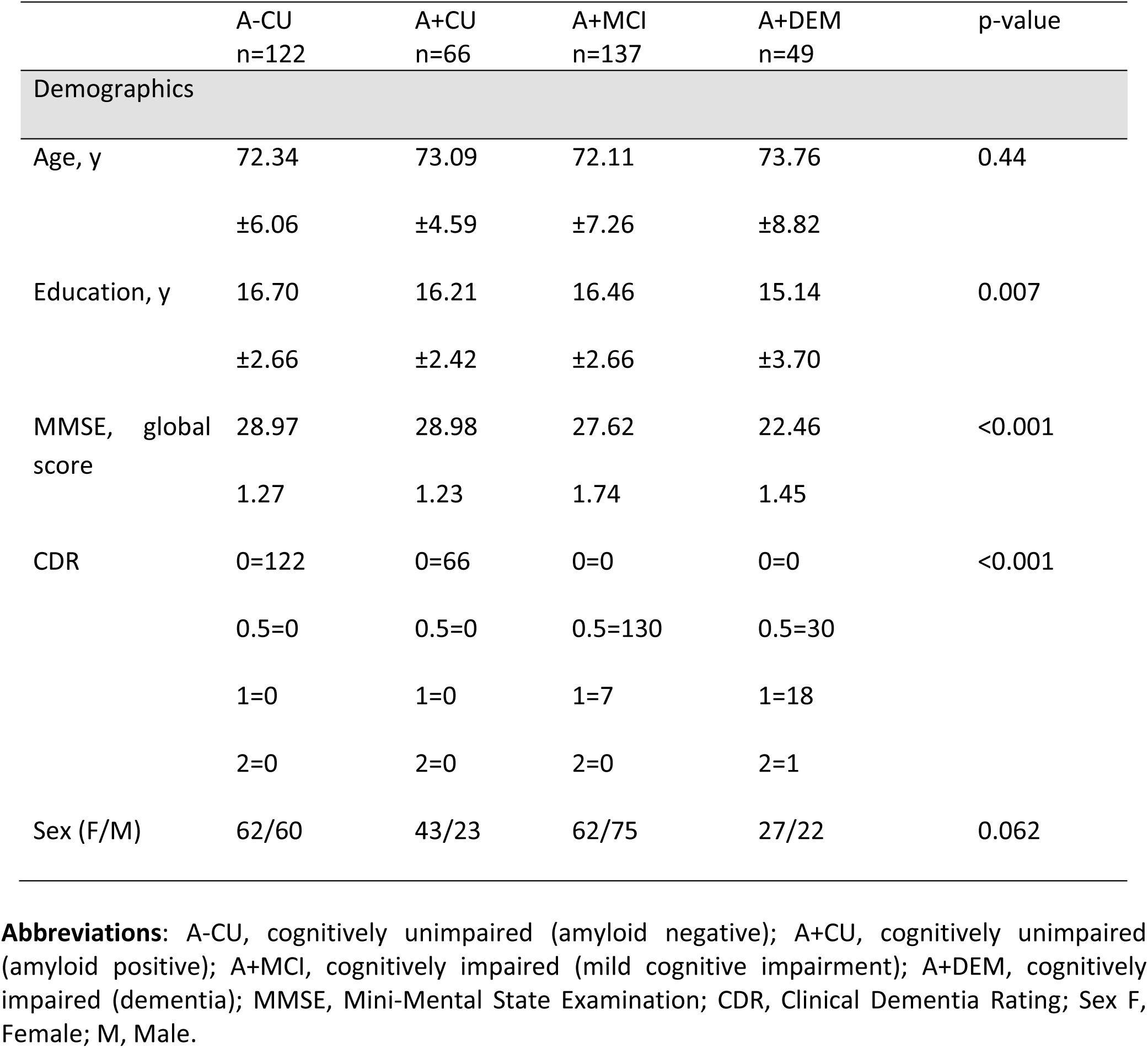
Baseline demographic characteristics of included participants.

**Table 2.** Baseline metabolic characteristics of included participants.

|  | A-CU<br>n=122 | A+CU<br>n=66 | A+MCI<br>n=137 | A+DEM<br>n=49 | p-value |
| --- | --- | --- | --- | --- | --- |
| Blood pressure and metabolic assessment |  |  |  |  |  |
| BMI | 27.14 | 26.02 | 26.00 | 25.24 | 0.004 |
|  | ±3.20 | ±3.56 | ±3.52 | ±3.43 |  |
| MAP | 94.04 | 99.08 | 95.79 | 96.68 | 0.009 |
|  | ±9.47 | ±11.15 | ±8.81 | ±11.29 |  |
| **TYG index** |  |  |  |  |  |
| TyG | 3.57 | 3.55 | 3.60 | 3.60 | 0.331 |
|  | ±0.19 | ±0.22 | ±0.19 | ±0.20 |  |

**Abbreviations:** A-CU, cognitively unimpaired (amyloid negative); A+CU, cognitively unimpaired (amyloid positive); A+MCI, cognitively impaired (mild cognitive impairment); A+DEM, cognitively impaired (dementia); BMI, body mass index; MAP, mean arterial pressure, TyG, triglyceride-glucose index.

**Table 3.**
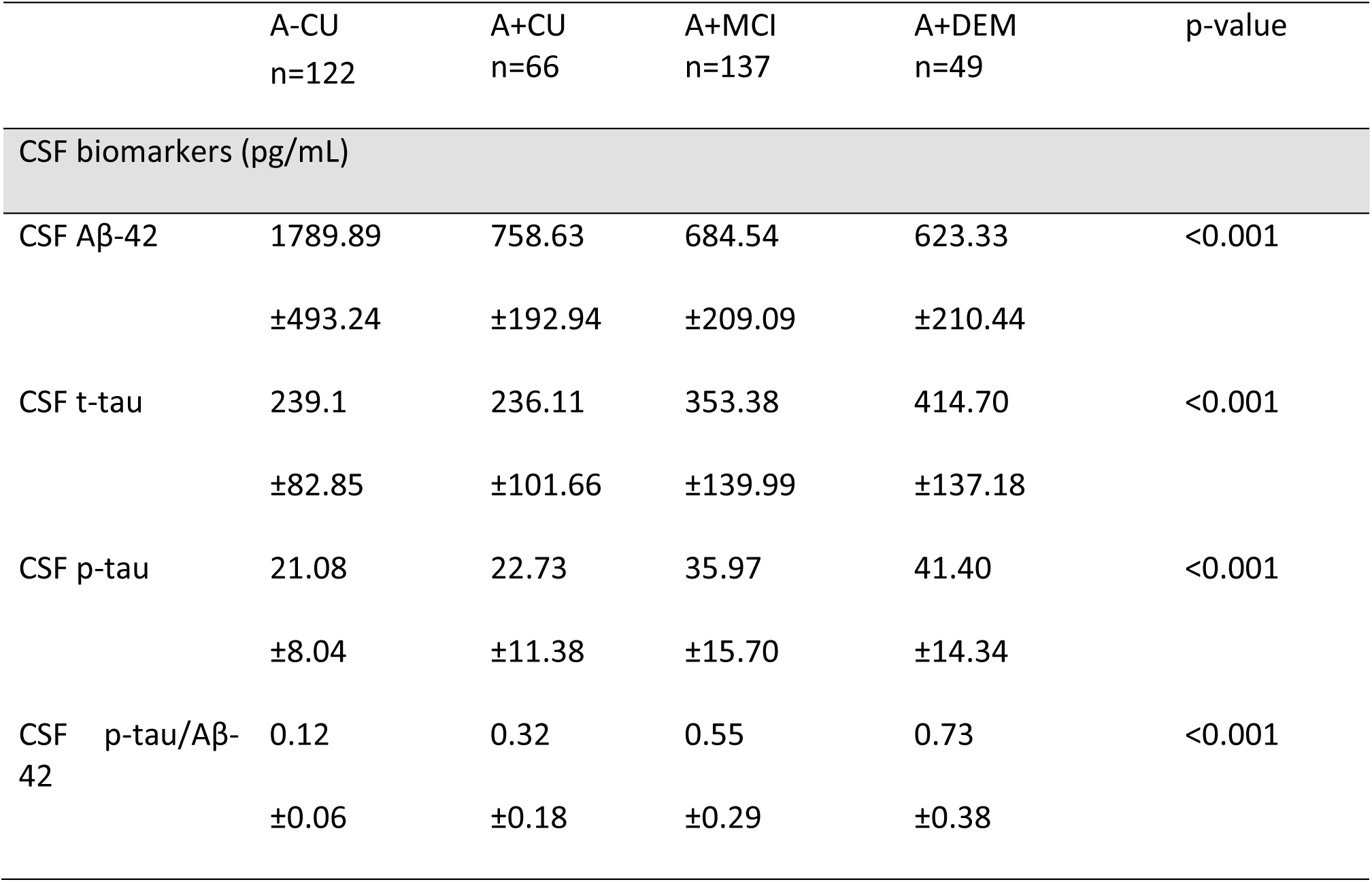
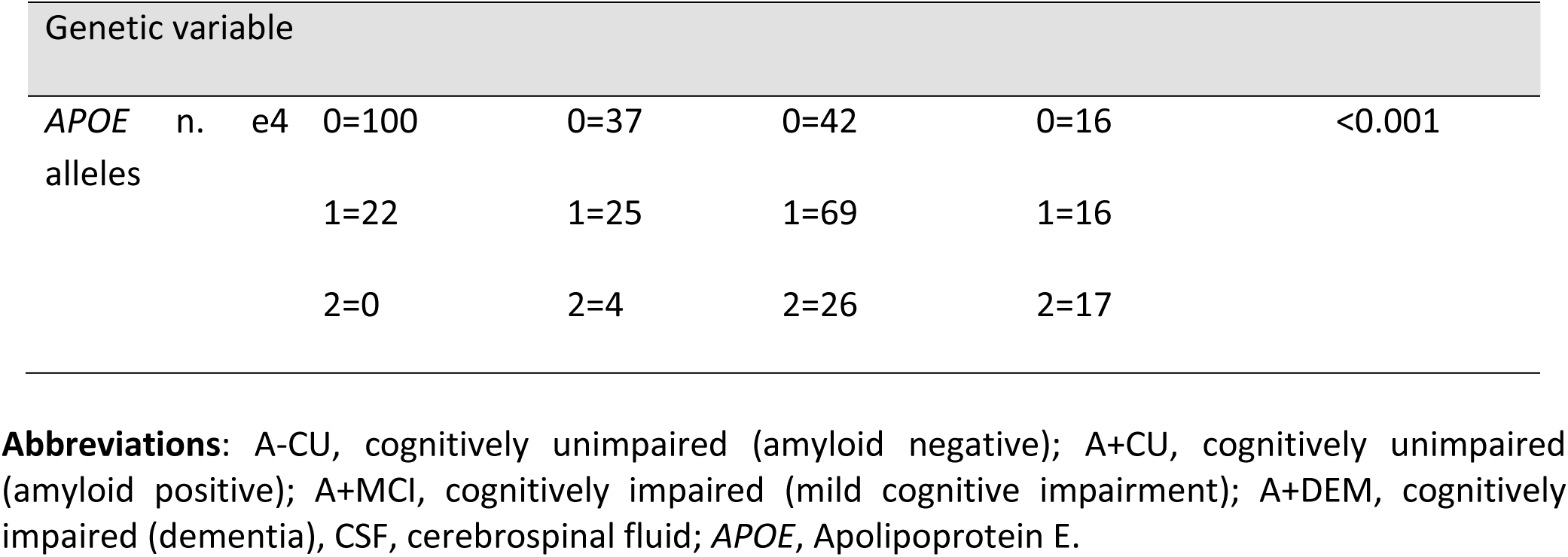
Baseline biological markers of included participants.

At baseline, no significant differences were observed across subgroups in age, sex, or Tyg levels. BMI (p=0.004) and education (p=0.007) increased along with increasing clinical severity. MAP displayed a distinct pattern across subgroups, with the highest levels observed in the A+CU group (p=0.009). According to methods, all demographics, MAP, BMI and *APOE* genotype were included in the final statistical analyses as confounding variables.

CSF biomarkers followed the subgroup definition, with increase in pathological levels with increase in clinical severity (p<0.001). Finally, *APOE* status also differed significantly across groups, with a progressively higher number of individuals carrying *APOE* ε4 alleles in A+MCI and A+DEM subgroups (p<0.001).

### Total sample – Voxel-based morphometry analysis

The initial multiple regression VBM analysis on the total sample revealed significant grey matter atrophy associated with AD diagnosis, particularly involving the temporal lobe, parietal lobe, limbic system, insular cortex, and cingulate cortex. These areas were subsequently selected for ROI-based analyses, adding to the total set of 36 regions of interest (Supplementary Table 2).

The voxel-wise interaction analysis between IR and diagnosis identified two significant clusters of grey matter volume, with different trends across diagnostic groups.

The first cluster, which survived false discovery rate (FDR) correction (pFDR = 0.032; kE = 1377), encompassed the right superior and middle temporal cortex.

The second cluster, which was significant at an uncorrected threshold (p-unc = 0.039; pFDR = 0.243; kE = 511), was observed in the bilateral anterior and middle cingulate cortex.

The post-hoc analyses on the two clusters revealed distinct patterns of neurodegeneration based on diagnosis. In cognitively unimpaired Aβ-negative individuals, no significant association was found between IR and grey matter volume. However, in cognitively unimpaired but Aβ-positive individuals, IR was linked to lower grey matter volume. This association was stronger in cognitively impaired individuals, with A+MCI showing a notable reduction in grey matter volume at high IR levels. The most pronounced effect was observed in A+DEM, where IR was associated with the greatest loss of grey matter volume (Figure 1).

**Figure 1.**
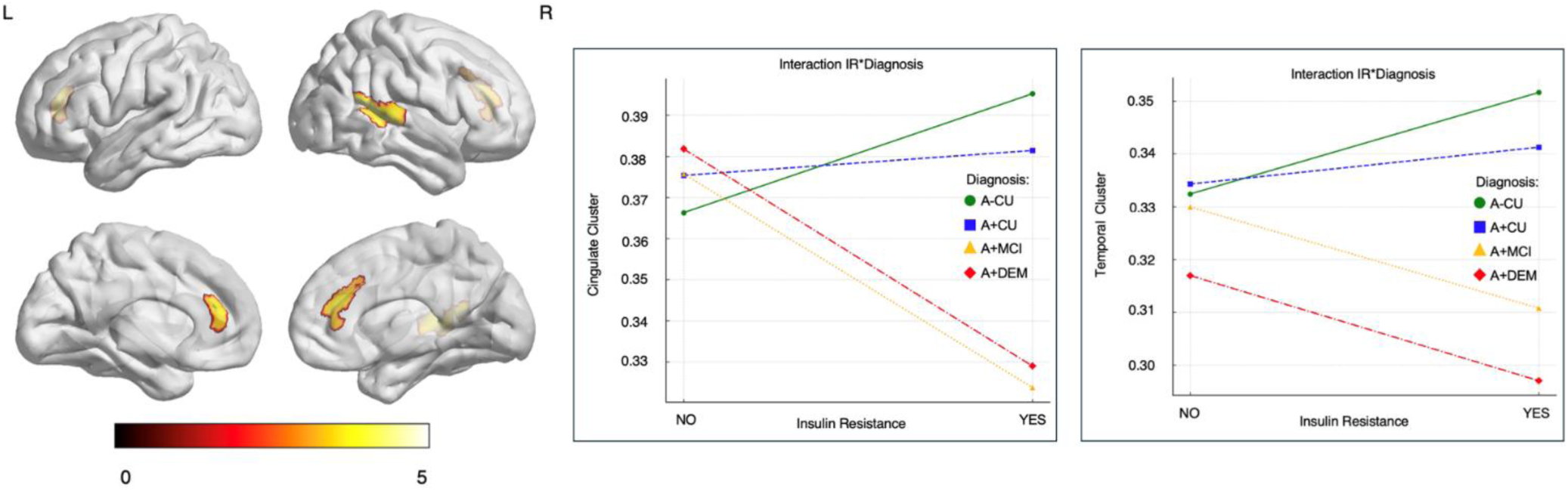
Total sample - voxel wise and post-hoc interaction analyses on two clusters showing different neurodegenerative trajectories across different stages of Alzheimer’s continuum.

### ROI-based analyses conducted within the total sample evidenced 12 significative regions of interest where the interaction effect of IR and diagnosis was significant

These areas included the bilateral temporal pole, bilateral superior temporal gyrus, bilateral parahippocampal gyrus, bilateral amygdala, right anterior and middle cingulate gyrus, left entorhinal area and left hippocampus. All these areas survived at FDR multiple comparisons correction.

The direction of the interaction effect was consistent with the previous voxel-wise and post-hoc analyses, where significant associations between high levels of IR and lower grey matter volume were detected only in subjects within the AD continuum from at-risk to symptomatic phases. In the parahippocampal gyrus and temporal pole, this effect was detected starting from the simple positivity to Aβ pathology A+CU, while in limbic areas, the effect was stronger in moderate/severe clinical stages (Supplementary Table 3).

### Subgroup analyses-CI subjects adjusted for disease severity

The voxel-wise multiple regression analysis of IR additionally adjusted for disease severity in the CI subgroup (A+MCI and A+DEM) identified three significant clusters of grey matter volume reductions.

The first cluster (pFDR = 0.002, kE = 2850) includes the bilateral anterior and middle cingulate gyrus, the right superior frontal gyrus medial segment and the bilateral supplementary motor cortex. The second cluster (pFDR = 0.026; kE = 1565) is located in the right hemisphere encompassing the hippocampus, parahippocampal gyrus, amygdala and superior temporal pole.

The third cluster (p-unc= 0.018; kE = 810) includes the left hemisphere, in particular the amygdala, parahippocampal gyrus, superior temporal pole, hippocampus and the inferior orbitofrontal cortex (Figure 2).

**Figure 2.**
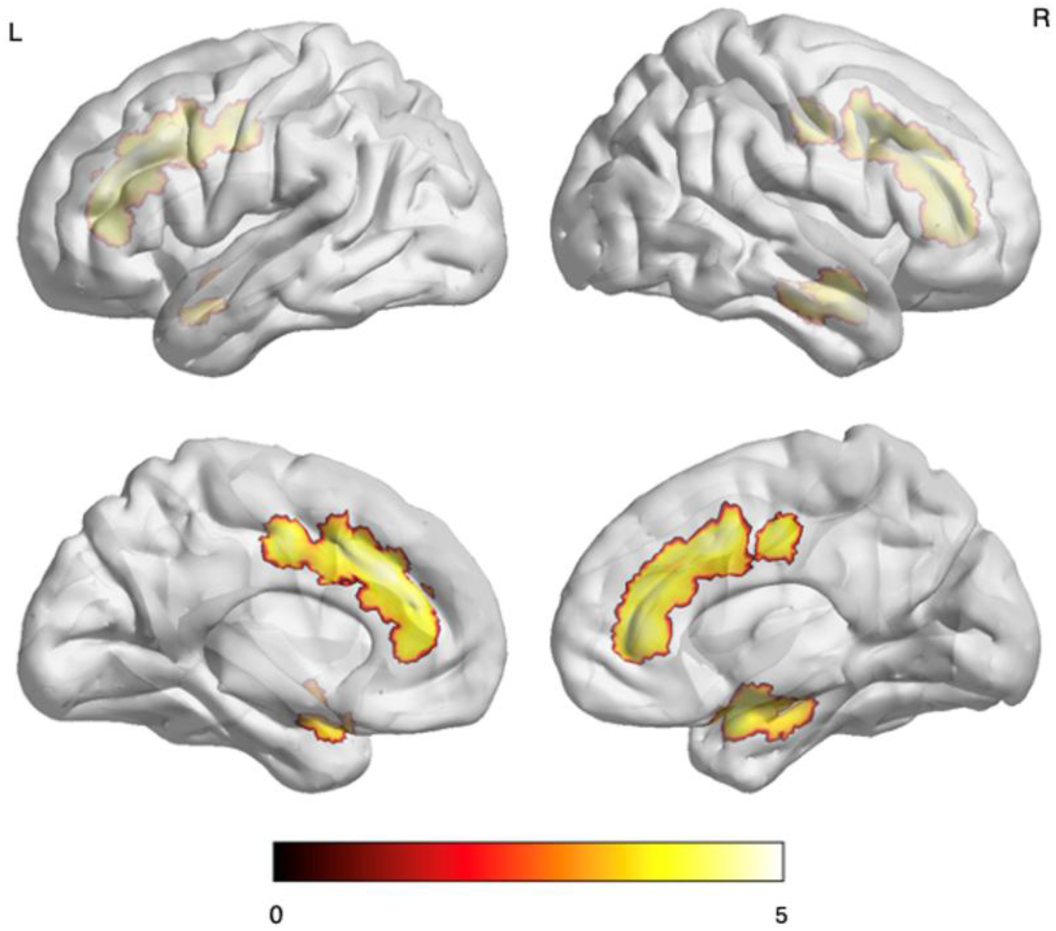
AD subgroup - voxel wise analysis, three clusters of grey matter reduction in association with insulin resistance.

ROI-based analyses conducted within the CI subgroup further adjusted for MMSE evidenced several areas surviving at FDR rate including limbic, insular, temporal, and parietal areas, with a predominance in the right hemisphere (Supplementary Table 4). Subgroup analyses-CU subjects

The voxel-wise interaction analysis between IR and Amyloid positivity identified one cluster, located in the left fusiform gyrus, inferior temporal gyrus and inferior temporal pole. This cluster only survived uncorrected and size thresholds (pFDR=0.183; kE=533; p-unc= 0.029).

The mean values from the detected cluster were extracted and then examined in a post-hoc analysis in order to further explore the directionality of this significant interaction.

In this analysis, interaction between IR and Amyloid suggests that IR is associated with grey matter volume changes in the amyloid-positive group, while this effect is absent in the amyloid-negative group (p<0.001) (Figure 3).

**Figure 3.**
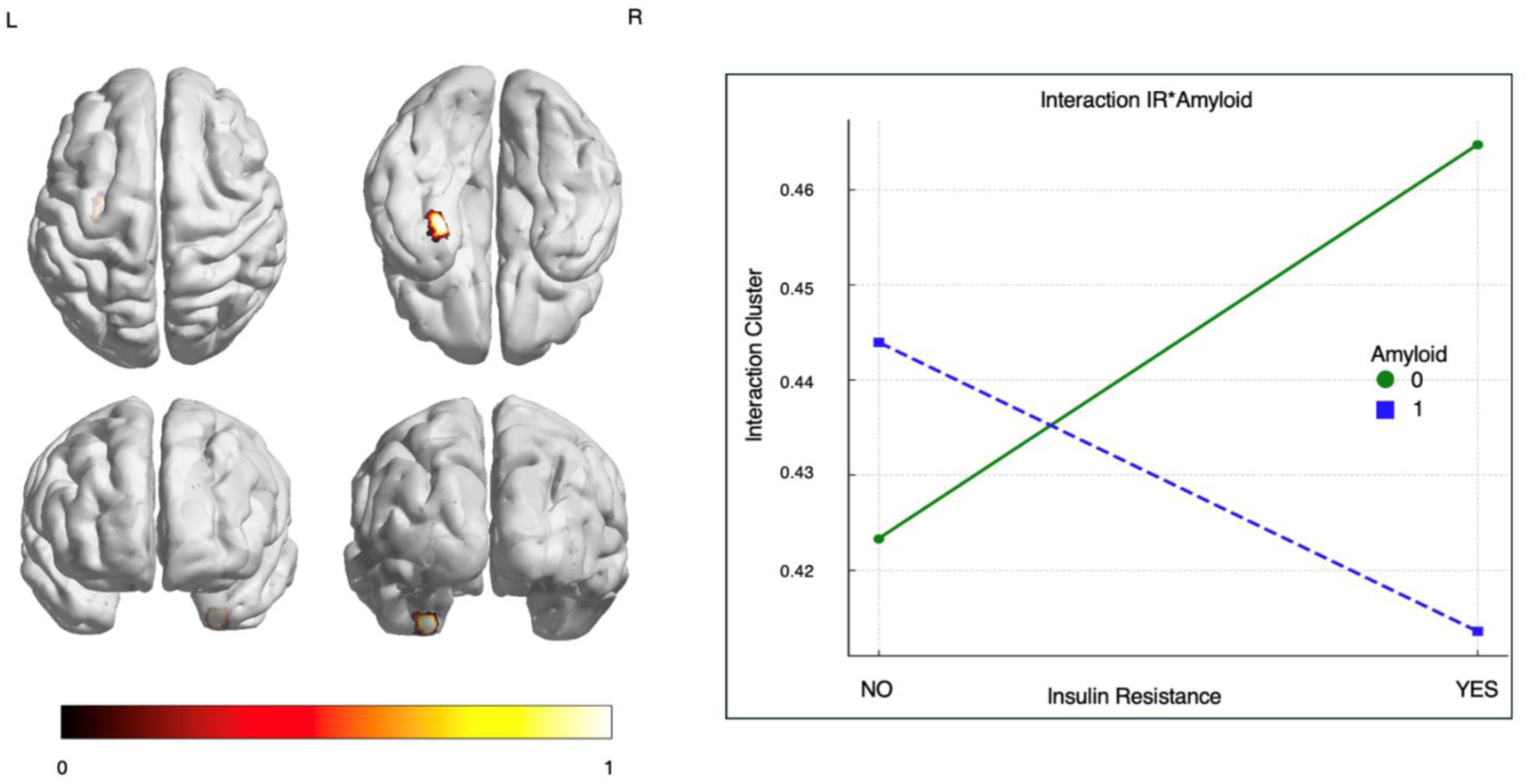
CU subgroup – voxel wise and post-hoc interaction analyses on the cluster showing different neurodegenerative trajectories between A+ and A-cognitively unimpaired individuals.

## Discussion

This study provides novel evidence stage-dependent relationship between IR and grey matter atrophy across the Alzheimer’s disease (AD) continuum. By combining voxel-based morphometry (VBM) and region-of-interest (ROI) analyses in a well-characterized cohort derived from the ADNI, we observed that insulin resistance may be associated with structural differences in brain regions known to be vulnerable to AD pathology. Notably, the deleterious effects of IR on grey matter were more pronounced in individuals with established amyloid pathology, while cognitively unimpaired, amyloid-negative individuals showed no significant structural alterations related to IR. Importantly, levels of the TyG index did not significantly differ across diagnostic groups, suggesting that the observed structural effects are not simply driven by group differences in metabolic status. These findings provide preliminary support for the hypothesis that metabolic dysfunction accelerates neurodegeneration in the presence of pathological AD substrates.

Previous studies have shown that IR is associated with increased brain vulnerability in cognitively healthy older adults. Our results evaluating the ADNI dataset demonstrated for the first time an interaction between clinical phases of AD and IR in driving structural vulnerability, independently from other known factors.

VBM analyses in the whole cohort revealed that IR-related atrophy localized to regions including the bilateral cingulate cortex, hippocampus, parahippocampal gyrus, and amygdala. These findings were reinforced by ROI-based analyses, which identified consistent IR-by-diagnosis interactions across 12 regions, all of which survived multiple comparison correction.

This evidenced a possible IR related region- and stage-dependent effect on neurodegeneration in AD continuum, highlighting its interaction with clinical course. Specifically, IR was observed to be associated with atrophy in specific brain regions, but its effects diminished in the most advanced stages of the disease, likely due to other drivers of neurodegeneration.

In the early phases of the disease, IR was associated with lower grey matter volumes in parahippocampal areas and the temporal pole, even in CU individuals who were amyloid-positive. This suggests that metabolic dysfunction increases the vulnerability of regions already susceptible to early AD pathology even in at-risk subjects, potentially opening an interesting window of intervention. This finding is consistent with the Braak and Braak staging model, which identifies the transentorhinal and parahippocampal cortices as the first cortical sites affected by AD pathology, i.e., Braak stages I-II^29,30^.

As the disease progresses into the early symptomatic stages (A+MCI), the effect of IR extends to the entorhinal cortex, to limbic regions like hippocampus and amygdala, and to other temporal regions, aligning with Braak stages III-IV, when tau pathology spreads into the limbic system. At this stage, IR is observed to be significatively associated with grey matter loss in these areas, supporting the hypothesis that metabolic dysfunction interacts with AD pathology to accelerate structural brain changes.

Of note, in A+DEM, IR no longer exerts a significant effect in these specific regions and even at whole-brain level, in the current cohort. This suggests that by this stage, extensive atrophy has already taken place, potentially involving other factors and leaving less impact of metabolic dysfunction on brain atrophy. This supports the idea that once a critical threshold of neurodegeneration is reached, the influence of modifiable risk factors such as IR becomes negligible or less relevant^31,32^.

An exception to this pattern is the cingulate cortex, where IR continues to exert an effect even A+DEM stages. This region showed vulnerability to IR starting from the MCI stage, and the effect persisted with a similar trend in the later clinical stages of the disease. This would argue for cingulate cortex remaining susceptible to metabolic dysfunction even when other brain regions have already undergone extensive atrophy. The cingulate cortex plays a key role in the default mode network (DMN), which is known to be disrupted early in AD and remains functionally compromised throughout disease progression^33–36^. Additionally, post-mortem studies confirm that the cingulate cortex is affected by both tau pathology (Braak V-VI) and amyloid accumulation in the later stages of AD^29,37^, potentially representing a further vulnerable region in later stages.

The study additionally examined the role of IR separately at different cognitive stages, in order to avoid a severity effect on grey matter integrity mainly driven by clinical severity. When analyzing the CI subgroup separately and thus additionally adjusting for cognitive severity (i.e., MMSE), IR was confirmed to play a relevant role on key AD-related brain areas independently from vascular, and demographics/clinical factors^38,39^.

In the analyses separately conducted on CU individuals, IR was found to already exhibit a neuropathological effect, albeit differently between amyloid-negative and amyloid-positive individuals. Notably, in amyloid-positive CU individuals, IR was significantly associated with lower grey matter volume in key regions, reinforcing the hypothesis that metabolic dysfunction may undermine the brain’s ability to cope with early AD pathology^40^. These findings are consistent with previous evidence suggesting that metabolic alterations, including reduced insulin and elevated HbA1c levels, are associated with early amyloid accumulation and neurodegeneration in CU, non-diabetic individuals^41^. Even in the absence of cognitive symptoms, IR appears to contribute to neuronal vulnerability, supporting the idea that it may play a role in the transition from preclinical to symptomatic AD^42–44^. This is supported by longitudinal evidence showing that IR measured in midlife is associated with increased amyloid deposition and cognitive decline in later life, even in the absence of cerebrovascular damage^45^.

These findings align with multi-level impact of IR on brain structural changes, in line with larger studies in aging population without amyloid stratification^46^. Of note, no interaction between *APOE* genotype and IR was observed in the cohort. One the one hand, this is an expected finding, as *APOE* appeared to play a major role in modulating the relationship between IR and BBB disruption. On the other hand, the limited number of homozygous *APOE* ε4 subjects could potentially explain these findings and larger cohorts in multicenter studies evaluating these specific subpopulation are warranted^47–49^.

It is crucial to underline that the impact of IR in our study was independent from other vascular risk factors, especially from obesity and hypertension. Obesity and hypertension are key factors associated with IR and, therefore, were included in all the analyses performed in the current study. This issue has several implications, as IR should be assessed and considered even in subjects with benign vascular profile. Subjects with IR are particularly suitable for both non-pharmacological and pharmacological interventions. Previous experimental studies have shown that antidiabetic drugs such as probucol and metformin may prevent cognitive deficits by attenuating the neuroinflammation and neurodegeneration mediated through BBB protective properties in dietary-induced pre-diabetic insulin-resistant mouse model. Thus, drugs currently approved to treat metabolic dysfunction hold promise to improve BBB function and reduce the pace of cognitive impairment. The glucagon-like peptide-1 (GLP-1) analogue liraglutide crosses the BBB and improves cognition in animal models while an exploratory trial of the GLP-1 analog dulaglutide found potential for slowing cognitive decline in T2DM patients^50^.

Addressing these risk factors in early clinical stages of AD may enhance the brain’s resilience against pathology. IR appears to weaken the brain’s ability to compensate for amyloid-driven damage, making metabolic intervention a promising avenue for neuroprotection, especially in at-risk and early clinical stages.

These approaches should be integrated with existing strategies aimed at increasing cognitive reserve, such as education, cognitive training, and social engagement, to provide a more comprehensive protective framework against AD. The assessment of IR in disease-modifying strategies will be also pivotal for a further understanding of its role as potential modulator of anti-amyloid effect potentially via BBB integrity alteration^13^. Overall, this study demonstrates the effect of IR on specific gray matter regions in a highly selected AD cohort and highlights the relevance of metabolic changes on the neurodegeneration along the clinical continuum of AD, with potential therapeutic implications.

## Author Contributions

V.B., A.G., and A.Pi. contributed to the analysis and interpretation of the data and drafted the manuscript. E.B., C.T., D.C., B.G., I.L., D.F., and T.F.O. critically revised the manuscript for important intellectual content. A.Pa. and A.Pi. were responsible for the design and conceptualization of the study, contributed to data interpretation, and revised the manuscript. All authors approved the final version of the manuscript and agree to be accountable for the accuracy and integrity of the work.

## Data Availability

The data used in this study are from the Alzheimer's Disease Neuroimaging Initiative (ADNI) and are available from the LONI data repository upon registration and data use agreement (https://adni.loni.usc.edu).

## Acknowledgement

Data collection and sharing for this project was funded by the Alzheimer’s Disease Neuroimaging Initiative (ADNI) (National Institutes of Health Grant U01 AG024904) and DOD ADNI (Department of Defense award number W81XWH-12-2-0012). ADNI is funded by the National Institute on Aging, the National Institute of Biomedical Imaging and Bioengineering, and through generous contributions from the following: AbbVie, Alzheimer’s Association; Alzheimer’s Drug Discovery Foundation; Araclon Biotech; BioClinica, Inc.; Biogen; Bristol-Myers Squibb Company; CereSpir, Inc.; Cogstate; Eisai Inc.; Elan Pharmaceuticals, Inc.; Eli Lilly and Company; EuroImmun; F. Hoffmann-La Roche Ltd and its affiliated company Genentech, Inc.; Fujirebio; GE Healthcare; IXICO Ltd.; Janssen Alzheimer Immunotherapy Research & Development, LLC.; Johnson & Johnson Pharmaceutical Research & Development LLC.; Lumosity; Lundbeck; Merck & Co., Inc.; Meso Scale Diagnostics, LLC.; NeuroRx Research; Neurotrack Technologies; Novartis Pharmaceuticals Corporation; Pfizer Inc.; Piramal Imaging; Servier; Takeda Pharmaceutical Company; and Transition Therapeutics. The Canadian Institutes of Health Research is providing funds to support ADNI clinical sites in Canada. Private sector contributions are facilitated by the Foundation for the National Institutes of Health (www.fnih.org). The grantee organization is the Northern California Institute for Research and Education, and the study is coordinated by the Alzheimer’s Therapeutic Research Institute at the University of Southern California. ADNI data are disseminated by the Laboratory for Neuro Imaging at the University of Southern California. CSF β-Amyloid42 biomarker positivity was defined using cut-offs established for the Elecsys® assays by Roche Diagnostics. The brain clusters were visualized with the BrainNet Viewer (http://www.nitrc.org/projects/bnv/) (Xia, M; Wang, J; He, Y. 2013). The study has been partially supported by #NEXTGENERATIONEU (NGEU) and funded by the Ministry of University and Research (MUR), National Recovery and Resilience Plan (NRRP), project MNESYS (PE0000006) – a multiscale integrated approach to the study of the nervous system in health and disease (DN. 1553 11.10.2022) – subproject DIGI-BRAIN and Funded by the European Union - Next Generation EU - NRRP M6C2 - Investment 2.1 Enhancement and strengthening of biomedical research in the NHS-PNRR – PNRR.

