## Supplementary material for "The Impact of Insulin Resistance on Grey Matter Changes Along the Alzheimer’s Disease Continuum Insulin Resistance and Grey Matter in AD"

**Supplementary figure 1.** Flowchart representing participants' enrollment and the final sample, with a subsequent division of cohorts.

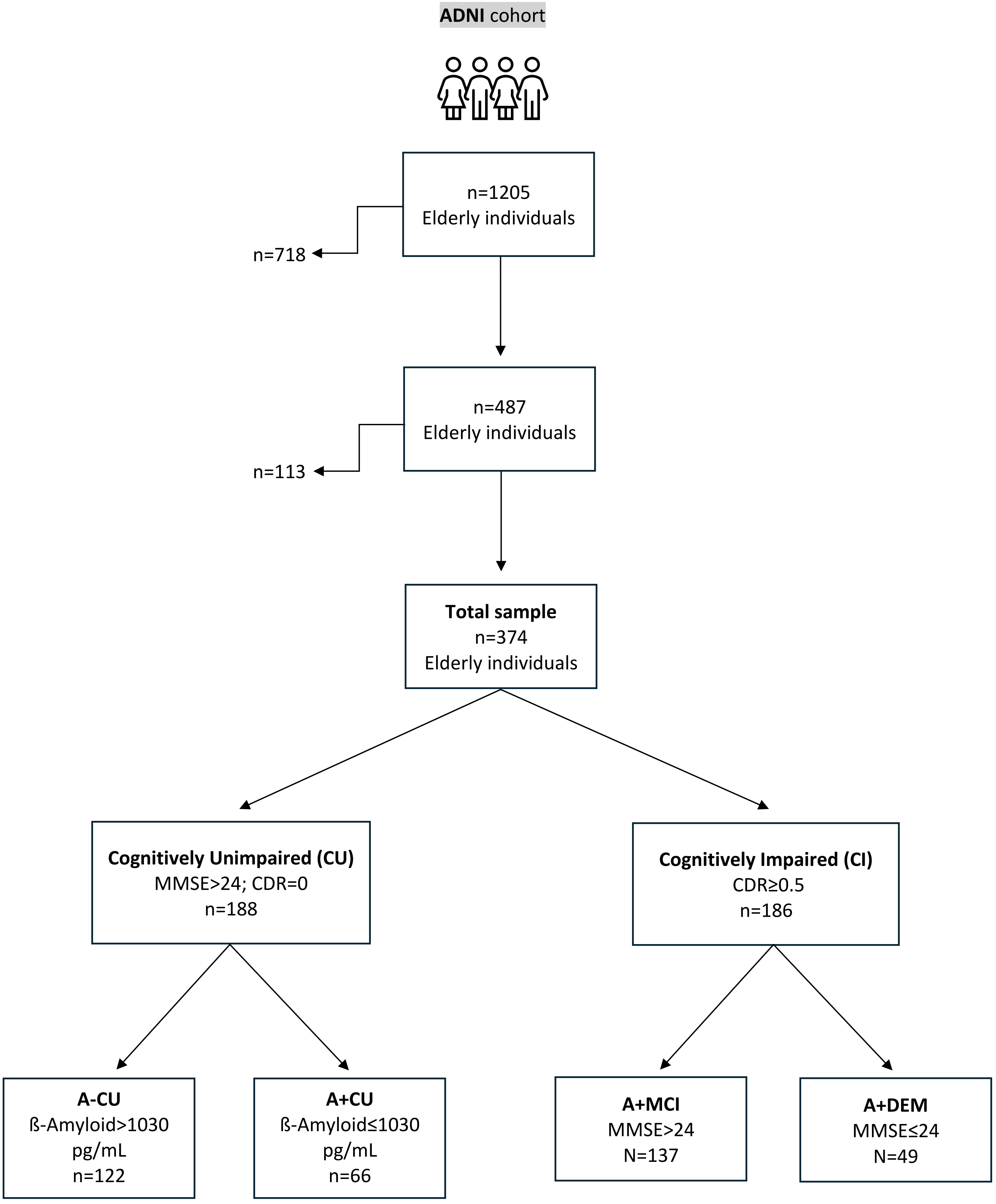

**Supplementary Table 1**. 18 bilateral Regions of Interest selected for ROI-based analyses.

| | |
|---|---|
| L/R transverse temporal gyrus | L/R precuneus |
| L/R temporal lobe | L/R superior parietal lobule |
| L/R superior temporal gyrus | L/R posterior insula |
| L/R planum temporale | L/R anterior insula |
| L/R middle temporal gyrus | L/R posterior cingulate gyrus |
| L/R inferior temporal gyrus | L/R anterior cingulate gyrus |
| L/R entorhinal area | L/R middle cingulate gyrus |
| L/R parahippocampal gyrus | L/R angular gyrus |
| L/R hippocampus | L/R amygdala |

**Supplementary Table 2**. Significant voxel-wise grey matter reductions in different cohorts - peak level significance.

| Cluster size k (mm³) | Region | MNI coordinates (mm) x | y | z | t-score | p-value |
|---|---|---|---|---|---|---|
| Total sample cohort n=374 | | | | | | |
| 1377 | Superior Temporal Cortex | 63 | -45 | 14 | 4.20 | <0.001 |
| | Middle Temporal Cortex | 68 | -34 | 4 | 4.12 | <0.001 |
| | Superior Temporal Cortex | 56 | -36 | 12 | 3.97 | <0.001 |
| 511 | Sup Anterior Cingulate Cortex L | 3 | 36 | 24 | 3.78 | <0.001 |
| | Middle Cingulate Cortex R | 14 | 36 | 30 | 3.72 | <0.001 |
| | Pre Anterior Cingulate Cortex L | 2 | 42 | 18 | 3.69 | <0.001 |
| CI cohort n=186 | | | | | | |
| 2850 | Middle Cingulate Cortex L | 2 | 27 | 33 | 4.72 | <0.001 |
| | Anterior Cingulate Cortex L | 2 | 39 | 22 | 4.44 | <0.001 |
| | Anterior Cingulate Cortex R | 3 | 15 | 39 | 4.40 | <0.001 |
| 1565 | Superior Temporal Pole R | 24 | 8 | -21 | 4.13 | <0.001 |
| | Hippocampus R | 12 | 0 | -16 | 3.96 | <0.001 |

| | | | | | | |
|---|---|---|---|---|---|---|
| | Parahippocampal Gyrus R | 21 | -10 | -16 | 3.67 | <0.001 |
| 810 | Insula L | -36 | 0 | -18 | 3.85 | <0.001 |
| | Orbitofrontal Cortex post L | -22 | 9 | -22 | 3.54 | <0.001 |
| | Parahippocampal Gyrus L | -14 | 2 | -20 | 3.23 | 0.001 |
| CU cohort n=188 | | | | | | |
| 553 | Fusiform L | -27 | -10 | -40 | 4.04 | <0.001 |

**Supplementary table 3**. ROI analyses on total sample – interaction and significance level.

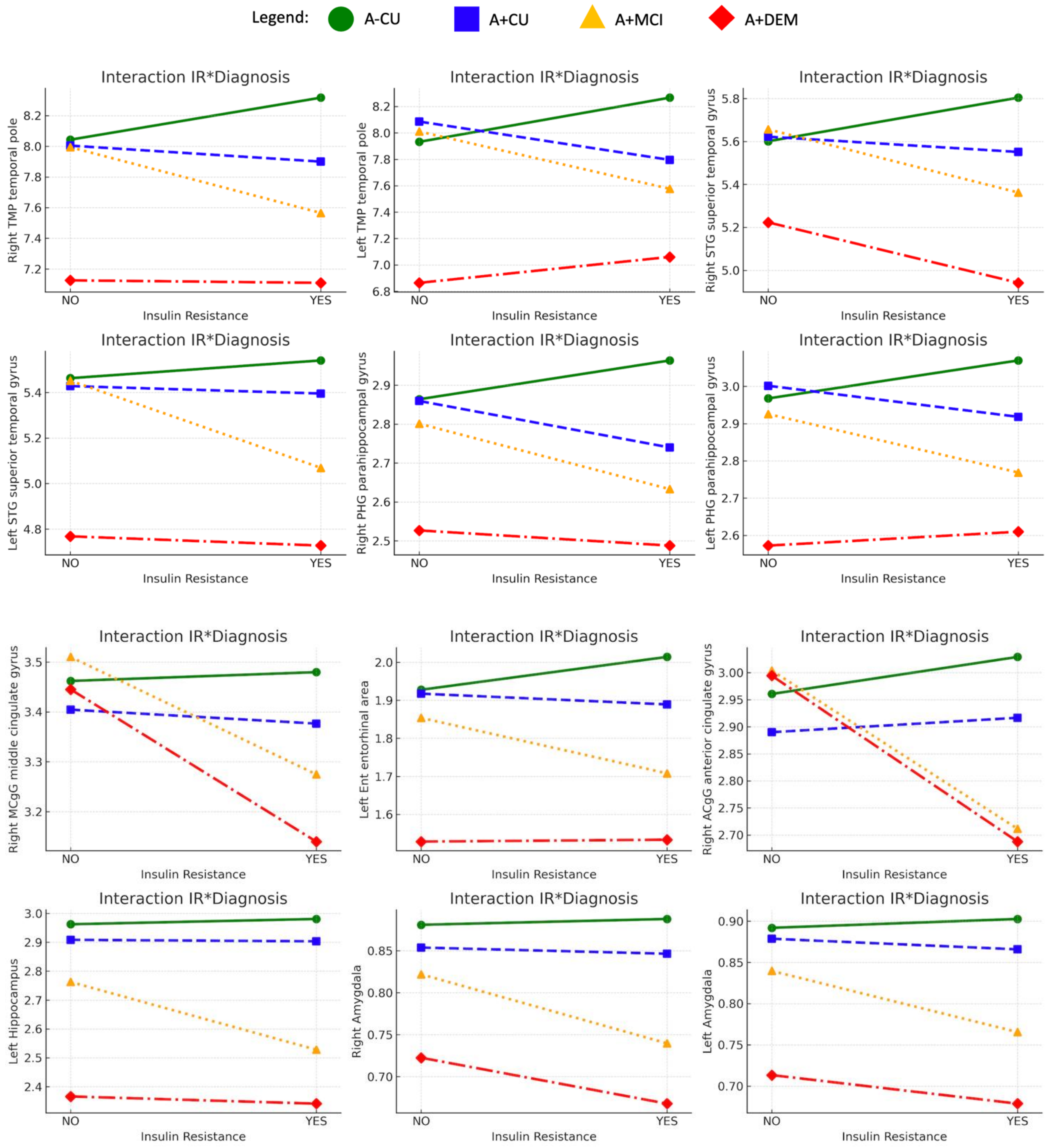

| ROI | P value uncorrected | pFDR |
|---|---|---|
| Right temporal pole | 0.044 | 0.044 |
| Left temporal pole | 0.005 | 0.019 |
| Right superior temporal gyrus | 0.015 | 0.020 |
| Left superior temporal gyrus | 0.022 | 0.024 |
| Left parahippocampal gyrus | 0.008 | 0.019 |
| Right parahippocampal gyrus | 0.010 | 0.019 |
| Right middle cingulate gyrus | 0.019 | 0.022 |
| Left entorhinal area | 0.007 | 0.019 |
| Right anterior cingulate gyrus | 0.004 | 0.019 |
| Left hippocampus | 0.008 | 0.019 |
| Left amygdala | 0.013 | 0.019 |
| Right amygdala | 0.012 | 0.019 |

**Supplementary Table 4**. ROI analyses on CI subgroup – significance level.

| ROI | P value uncorrected | pFDR |
|---|---|---|
| Right transverse temporal gyrus | 0.003 | 0.006 |
| Right hippocampus | 0.001 | 0.003 |
| Right anterior cingulate gyrus | 0.001 | 0.001 |
| Right middle cingulate gyrus | 0.001 | 0.003 |
| Right amygdala | 0.001 | 0.003 |
| Right entorhinal area | 0.001 | 0.003 |
| Left amygdala | 0.001 | 0.003 |
| Left hippocampus | 0.001 | 0.003 |
| Right angular gyrus | 0.002 | 0.005 |
| Right anterior insula | 0.004 | 0.008 |
| Right planum polare | 0.005 | 0.009 |
| Right posterior insula | 0.001 | 0.003 |

| | | |
|---|---|---|
| Right precuneus | 0.011 | 0.015 |
| Right superior parietal lobule | 0.002 | 0.005 |
| Left superior temporal gyrus | 0.006 | 0.010 |
| Right posterior cingulate gyrus | 0.012 | 0.015 |
| Right superior temporal gyrus | 0.007 | 0.010 |
| Left angular gyrus | 0.037 | 0.037 |
| Right parahippocampal gyrus | 0.029 | 0.031 |
| Left entorhinal gyrus | 0.017 | 0.019 |
| Left posterior insula | 0.007 | 0.010 |
| Left anterior insula | 0.015 | 0.017 |
| Left planum polare | 0.012 | 0.015 |
| Right planum polare | 0.006 | 0.010 |
| Right middle temporal gyrus | 0.031 | 0.032 |
